# Comparison of valvular and ventricular function after right ventricular, leadless and left bundle branch area pacemakers

**DOI:** 10.1101/2025.05.10.25327288

**Authors:** Anja Zhou, Isabelle Andersson, Mutaz Alkalbani, Abbas Emaminia, Eunice Yang, Martin Ugander, Brett D Atwater

**Affiliations:** Department of Clinical Physiology, Karolinska University Hospital, and Karolinska Institute, Stockholm, Sweden; Inova Schar Heart and Vascular, Falls Church, Virginia, USA; Kolling Institute, Royal North Shore Hospital, and University of Sydney, St Leonard’s, Sydney, Australia

**Keywords:** cardiac pacemakers, echocardiography, tricuspid regurgitation, mitral regurgitation, left ventricular ejection fraction

## Abstract

**Introduction:** Changes in tricuspid regurgitation (TR), mitral regurgitation (MR) and left ventricular ejection fraction (LVEF) are frequently noted after conventional pacemaker implantation but prior studies evaluating whether left bundle branch area (LBBA) or leadless pacemakers modify those observed changes are limited. This study aims to compare changes in TR, MR and LVEF after implantation of conventional right ventricular (RV), leadless, and LBBA pacemakers.

**Methods:** Inclusion criteria were first-time pacemaker implantation and pre- and post-implant echocardiography. Change in TR, MR and LVEF were analyzed using post-hoc adjusted Kruskal-Wallis and chi-squared testing, and multivariable ordinal logistic regression.

**Results:** Among 400 consecutive patients (RV, n=228; LBBA, n=136; leadless, n=36), the change in TR grade differed between pacemaker types (median [interquartile range] grade change: LBBA 0[0,0], leadless 0[0,1], RV 0[0,1]; p<0.01). The prevalence of severe TR was similar between pacemaker groups before implant (p=0.93), but more frequent following implant of RV and leadless pacemakers compared to LBBA pacemakers (p=0.02). In multivariable ordinal logistic regression, leadless (OR 2.26, p=0.03) and RV pacemakers (OR 1.66, p=0.03) both predicted TR worsening compared to LBBA. The change in MR grade differed between pacemaker types (grade change: LBBA 0[-1,0], leadless 0[0,1], RV 0[0,0]; p<0.01). The change in LVEF differed between pacemaker types (LVEF change: LBBA 0[- 5,7]%, leadless -5[-14,1]%, RV -3[-9,2]%; p<0.01).

**Conclusion:** Change in TR, MR grade and LVEF following pacemaker implant varied by pacemaker type. Compared to leadless and RV, LBBA pacemaker implant was associated with more favorable changes in valvular and ventricular function.

**Graphical Abstract:** 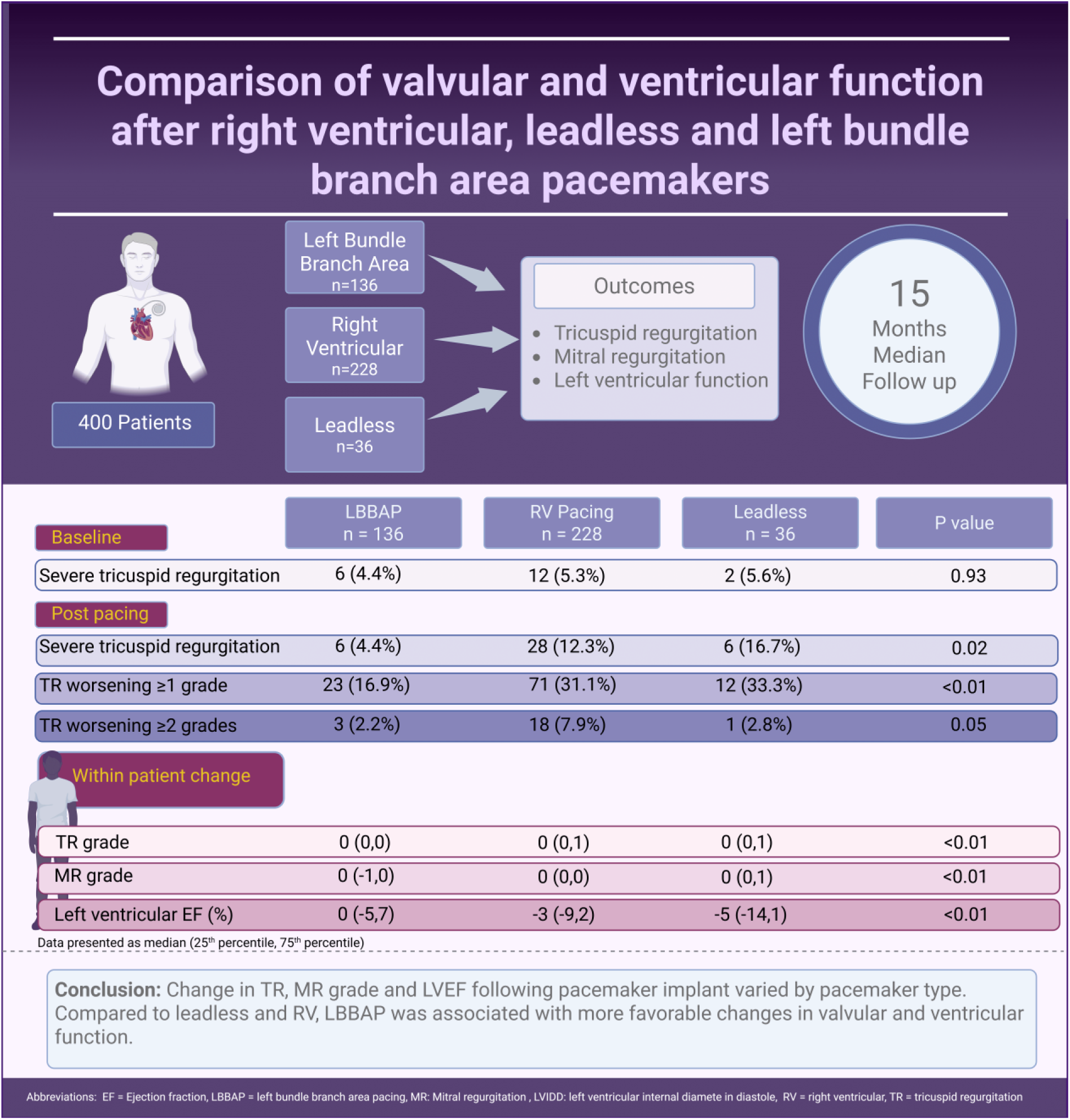

## Introduction

The use of permanent pacemakers is widespread and continues to increase in the United States (1). Pacemakers are the most frequently utilized cardiac implantable electronic device (CIED), and have been associated with worsening tricuspid regurgitation (TR) (2). CIED-related TR is associated with higher rates of heart failure hospitalization and all-cause and cardiovascular death (3, 4). There are several possible mechanisms behind CIED-related TR, including mechanical perforation and impingement of valve leaflets, subvalvular entanglement, and right ventricular (RV) septal to lateral wall dyssynchrony induced by continuous RV pacing resulting in annular dilation that impedes valvular coaptation (5). Conventional RV apical pacemakers can cause left ventricular (LV) dilation, reduced left ventricular ejection fraction (LVEF) and functional mitral regurgitation (MR) (6, 7). Recently, there has been an evolution in pacing mechanisms and devices. Conduction system pacing (CSP), encompassing His bundle pacing (HBP) and left bundle branch area (LBBA) pacing utilizes inherent conduction pathways and as such can maintain physiologic ventricular activation and function (7). Leadless pacemakers were developed to reduce lead and pocket complications and may reduce risk of mechanical tricuspid valve interference (8, 9). However, the findings of previous studies investigating TR after pacemaker implantation have been contradictory (5). A recent meta-analysis evaluating the risk of TR after pacemaker implant pooled patients with LBBA and HBP into a single CSP group making it difficult to discern whether LBBA reduces the risk of TR compared to other implant methods (4). This study aimed to compare changes in TR and MR severity, and LVEF, following implantation of conventional transvenous RV, leadless RV, and LBBA pacemakers.

## Methods

### Study population and data collection

Consecutive patients who received a *de novo* conventional RV pacemaker, LBBA pacemaker, or leadless pacemaker in the Inova Health System between 2017 and 2023 were retrospectively enrolled (Figure 1). Inclusion criteria were implantation of one of the specified pacemaker types and a pre-procedural echocardiogram performed ≤1 year before, and a post-procedural echocardiogram performed ≥3 months after implantation. Exclusion criteria were congenital heart disorders, primary pulmonary hypertension, previous tricuspid valve (TV) intervention, TV endocarditis, or heart transplant, chronic dialysis, pulmonary embolism between pre- and post-implant echocardiography, or poor or incomplete imaging precluding assessment of TR severity. Baseline data, pacing indication and ventricular pacing percentage were collected from electronic medical records.

**Figure 1.**
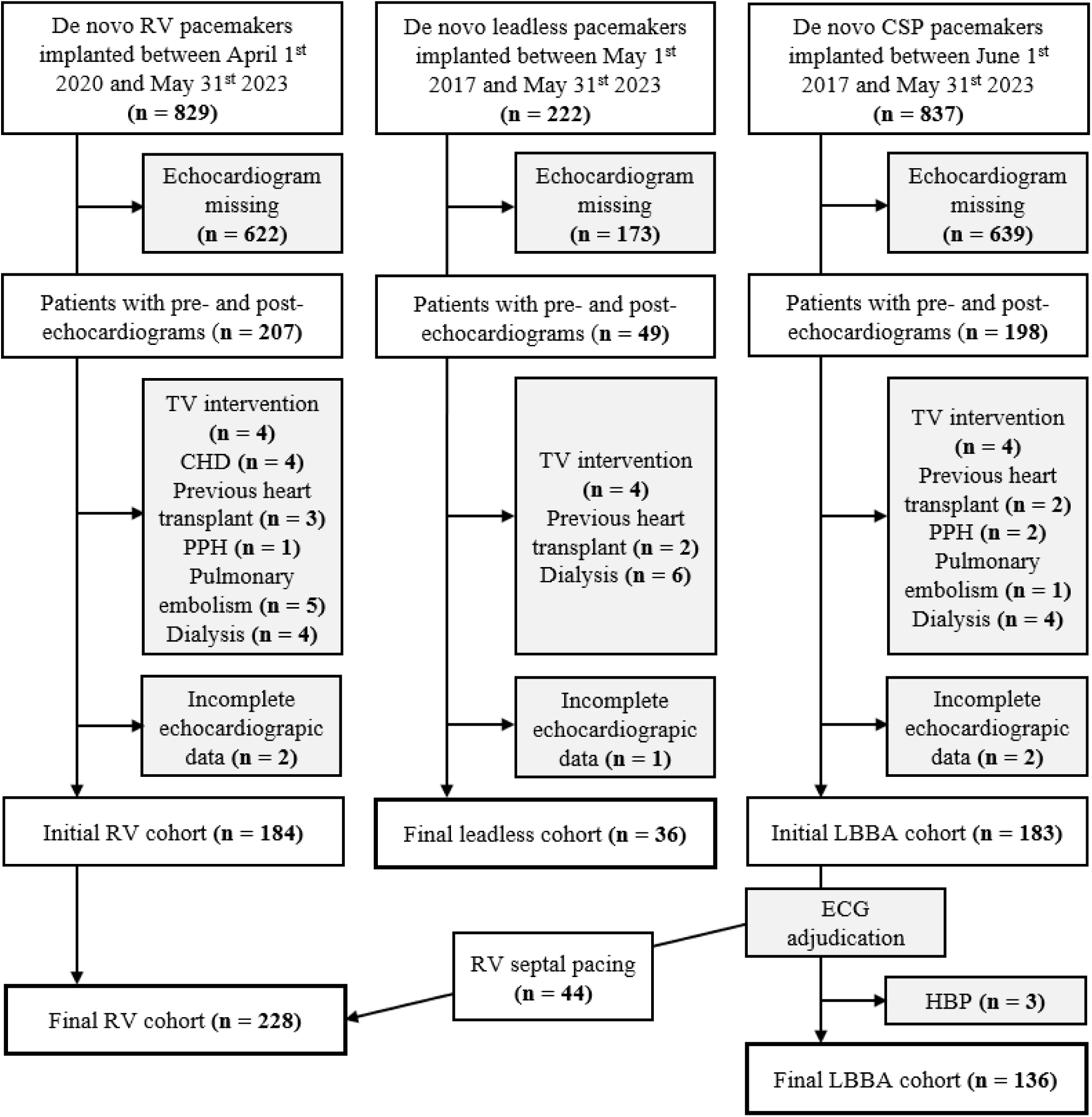
Patient selection. Abbreviations: CHD = congenital heart disorder, CSP = conduction system pacing, ECG = electrocardiogram, HBP = His bundle pacing, LBBA = left bundle branch area, PPH = primary pulmonary hypertension, RV = right ventricular, TV = tricuspid valve

### Device implantation

Board certified electrophysiologists implanted the devices in accordance with procedure guideline (10). LBBA pacemaker implantation was performed using a model 3830 lead (4.1 Fr, SelectSecure™, Medtronic, Minneapolis, MN) through a pre-formed sheath (model 315His or 304His, Medtronic, Minneapolis, MN) in the LBBA using fluoroscopic imaging and electrocardiogram (ECG) analyses. Leadless pacemakers (Micra™, Medtronic, Minneapolis, MN) were positioned in an apical or septal RV position using contrast injection and multiple fluoroscopic projections. RV pacing leads were implanted in the RV apex or septum using fluoroscopic imaging and manually curved or pre-formed stylets or in the RV septum through a pre-formed sheath. The security of fixation, position, and performance for all devices were ensured with post procedure anterior/posterior or anterior/posterior and lateral chest x rays and post procedure remote monitoring.

### Pacing group assignment

Left bundle branch capture (LBBC) was defined using criteria included in the EHRA/APHRS/CHRS/LAHRS and HRS/APHRS/LAHRS clinical consensus statements on conduction system pacing (10, 11). Specifically, LBBC included the presence of an R’ in lead V1 with an R-wave peak time ≤ 80ms in patients with an underlying wide QRS or ≤ 75ms in patients with narrow QRS. LV septal capture included patients with an R’ in lead V1 but an R-wave peak time exceeding criteria for LBBC. Patients meeting LBBC or LV septal capture definitions were considered to have LBBA pacing. Patients with an apical lead placement or a septal lead placement without an R’ in V1 (QS pattern) during unipolar pacing were included in the conventional RV pacing group.

### Echocardiographic assessment

TR severity was assessed from transthoracic echocardiograms by two of the authors (AZ, AE), according to an ordinal scale (0 = no regurgitation, 1 = mild regurgitation, 2 = moderate regurgitation, 3 = severe regurgitation). The assessment was made visually with color-, and continuous wave Doppler, assessing vena contracta, jet area, velocity curve density and contour, and systolic hepatic vein flow reversal. The post-procedural echocardiogram with the highest TR grade and longest follow-up was evaluated. Other echocardiographic measurements were determined by board-certified cardiologists in conjunction with clinical care. Those include mitral regurgitation (MR) grade, left ventricular ejection fraction (LVEF) calculated by the modified Simpson’s biplane method, tricuspid annular plane systolic excursion (TAPSE), left ventricular internal diameter in diastole (LVIDd), and right ventricular internal diameter in diastole (RVIDd). MR was graded according to the same ordinal scale as TR, with mild including trace, moderate including mild-to-moderate, and severe including moderate-to-severe. All echocardiographic assessments were done according to American Society of Echocardiography guidelines (12).

### Statistical analysis

Continuous data were presented as mean ± SD or median [interquartile range] depending on the normality of distribution. Normality was tested using the Shapiro-Wilk test. Categorical variables were presented with numbers and percentages. Group level analyses compared measurements before and after implantation using the paired t-test or paired Wilcoxon signed-rank test for continuous variables, and the chi-squared test for ordinal variables. Within patient change analyses compared differences between pacemaker groups using one-way analysis of variance (ANOVA) or Kruskal-Wallis testing for continuous and ordinal variables, with post-hoc Tukey or Dwass-Steel-Critchlow-Flingner adjustment for multiple comparisons. Frequency distributions were compared between pacemaker groups using the chi-squared test, with Bonferroni correction for multiple comparisons.

Univariable ordinal logistic regression was used to evaluate predictors of change in TR grade and MR grade. Variables evaluated through univariable screening included age, sex, body surface area, hypertension, coronary artery disease, heart failure, atrial arrythmias, chronic obstructive pulmonary disease, diabetes type 2, renal dysfunction, sinus node dysfunction, atrioventricular block and cardiac resynchronization therapy indication, use of angiotensin-converting enzyme inhibitors/angiotensin receptor blockers/angiotensin receptor-neprilysin inhibitors, beta blockers, sodium-glucose cotransporter-2 inhibitors, mineralocorticoid receptor antagonists and loop diuretics, ventricular pacing percentage, follow-up time, pacemaker type and TR grade, MR grade, LVEF, LVIDd, RVIDd and TAPSE at baseline. Multivariable ordinal logistic regression examined independent predictors of change in TR grade and MR grade incorporating variables with a p<0.1 in univariable analyses. All analyses were performed using Jamovi software (version 2.3.2), and statistical significance was considered at a two-sided p-value of <0.05.

## Results

### Baseline patient characteristics, echocardiographic data and device specifications

400 patients formed the final cohort in this study. 228 patients received a conventional RV pacemaker (185 with conventional stylet driven RV placement and 43 with catheter delivered deep septal RV placement), 36 patients received a RV leadless pacemaker, and 136 patients received a LBBA pacemaker (123 with LBBC and 13 with LV septal capture). Baseline characteristics, echocardiographic data and device specifications are presented in Table 1. There were no differences in age, sex, or weight across pacemaker groups. Atrial arrhythmias were more common in the leadless group while heart failure was more common in the LBBA group. Atrioventricular block (AVB) was the most common pacemaker indication for all pacemaker types. Cardiac resynchronization therapy (CRT) indication was only present in the LBBA group. Before implantation, the baseline TR and MR grade did not differ between pacemaker groups (p=0.25 and p=0.82 retrospectively), and the prevalence of severe TR was similar across all pacemaker groups (p=0.93). Pre-implant LVEF was highest in the RV group (p<0.01) and ventricular pacing burden was highest in the LBBA group (p<0.01). Follow-up duration was longer in the RV and leadless groups compared to the LBBA group (p<0.01).

**Table 1.** Baseline patient characteristics, echocardiographic data and device specifications.

|  | **RV (n = 228)** | **Leadless (n = 36)** | **LBBA (n = 136)** | **p-value** |
| --- | --- | --- | --- | --- |
| *Demographics* |  |  |  |  |
| Age, years | 79 [73, 84] | 82 [74, 89] | 78 [73, 83] | 0.07 |
| Female, n (%) | 102 (45) | 14 (39) | 57 (42) | 0.75 |
| Weight (kg) | 80.3 [70.3, 94.3] | 81.5 [63.1, 115] | 82.5 [70.5, 93.0] | 0.93 |
| Body surface area (m<sup>2</sup>) | 1.91 [1.76, 2.11] | 1.98 [1.71, 2.27] | 1.95 [1.76, 2.10] | 0.73 |
| *Cardiovascular disease history, n (%)* |  |  |  |  |
| Hypertension | 123 (54) | 22 (61) | 81 (60) | 0.49 |
| CAD | 67 (29) | 15 (42) | 34 (25) | 0.14 |
| Heart failure | 38 (17) | 8 (22) | 40 (29) | **0.02\*** |
| Atrial arrhythmias | 83 (36) | 24 (67) | 54 (40) | **<0.01\*** |
| *Comorbidities, n (%)* |  |  |  |  |
| COPD | 9 (4) | 2 (6) | 10 (7) | 0.37 |
| Diabetes type 2 | 48 (21) | 11 (31) | 29 (21) | 0.43 |
| Renal dysfunction<br>(eGFR < 60 ml/min/1.73 m<sup>2</sup>) | 91 (40) | 11 (31) | 52 (39) | 0.56 |
| *Pacemaker indication, n (%)* |  |  |  |  |
| SND | 96 (42) | 10 (28) | 38 (28) | **0.01\*** |
| AVB | 132 (58) | 26 (72) | 84 (62) | 0.25 |
| CRT | 0 (0) | 0 (0) | 14 (10) | **<0.01\*** |
| *Cardiovascular medications, n (%)* |  |  |  |  |
| ACE inhibitors/ARB/ARNI | 103 (45) | 16 (44) | 66 (49) | 0.80 |
| Beta blockers | 46 (20) | 7 (19) | 37 (27) | 0.27 |
| SGLT2 inhibitors | 2 (1) | 0 (0) | 10 (7) | **<0.01\*** |
| MRA | 17 (7) | 1 (3) | 22 (16) | **<0.01\*** |
| Loop diuretics | 72 (32) | 16 (44) | 53 (39) | 0.17 |
| *Echocardiographic and device data* |  |  |  |  |
| Baseline TR (grade) | 1 [1, 1] | 1 [0, 1] | 1 [0, 1] | 0.25 |
| Baseline severe TR, n (%) | 12 (5.3) | 2 (5.6) | 6 (4.4) | 0.93 |
| Baseline MR (grade) | 1 [1, 1] | 1 [1, 1] | 1 [1, 1] | 0.82 |
| Baseline LVEF, % | 62.5 [58.0, 66.0] | 61.0 [57.1, 65.0] | 59.5 [51.0, 64.0] | **<0.01\*** |
| Ventricular pacing burden, % | 49.5 [1.0, 98.1] | 55.5 [16.0, 91.4] | 98.4 [45.7, 99.7] | **<0.01\*** |
| Follow-up duration, months | 18.9 [11.9, 27.1] | 14.1 [9.8, 19.9] | 11.8 [8.0, 17.1] | **<0.01\*** |

Data presented as median [25<sup>th</sup> percentile, 75<sup>th</sup> percentile] or number, n (%). Regurgitation grade: none = 0, mild = 1, moderate = 2, severe = 3. Comparisons between groups were made using the chi-squared or Kruskal-Wallis test. \* p<0.05
Abbreviations: ACE = angiotensin-converting enzyme, ARB = angiotensin receptor blockers, ARNI = angiotensin receptor-neprilysin inhibitors, AVB = atrioventricular block, CAD = coronary artery disease, COPD = chronic obstructive pulmonary disease, CRT = cardiac resynchronization therapy, eGFR = estimated glomerular filtration rate, LBBA = left bundle branch area, LVEF = left ventricular ejection fraction, MRA = mineralocorticoid receptor antagonists, RV = right ventricular, SGLT2 = sodium-glucose cotransporter-2, SND = sinus node dysfunction, TR = tricuspid regurgitation

### Changes in tricuspid valve function

#### Group level changes

After a median follow-up of 15.4 [10.0, 23.9] months, the overall TR grade in the entire cohort increased after pacemaker implantation (TR grade pre: 1 [0, 1], TR grade post: 1 [1, 1]; p<0.01). Group level analyses comparing the echocardiographic measures before and after each pacemaker type are shown in Table 2. The frequency distribution of TR grade severity was different pre-implant compared to post-implant in all pacemaker groups (RV, p<0.01; leadless, p<0.01; LBBA, p<0.01).

**Table 2.** Group level echocardiographic measurements compared before and after pacemaker implantation.

|  | RV (n = 228) |  |  |  |  | Leadless (n = 36) |  |  |  |  | LBBA (n = 136) |  |  |  |  |
| --- | --- | --- | --- | --- | --- | --- | --- | --- | --- | --- | --- | --- | --- | --- | --- |
|  | Post TR Severity |  |  |  |  | Post TR Severity |  |  |  |  | Post TR Severity |  |  |  |  |
| Pre-TR Severity | None | Mild | Moderate | Severe | p-value | None | Mild | Moderate | Severe | p-value | None | Mild | Moderate | Severe | p-value |
| None | 17(7.5) | 32(14) | 2(0.9) | 0(0) | <0.01* | 9(25) | 6(16.7) | 0(0) | 0(0) | <0.01* | 22(16.2) | 12(8.8) | 1(0.7) | 0(0) | <0.01* |
| Mild | 25(11) | 86(37.7) | 15(6.6) | 16(7) |  | 0(0) | 12(33.3) | 2(5.6) | 1(2.8) |  | 17(12.5) | 52(38.2) | 7(5.1) | 2(1.5) |  |
| Moderate | 0(0) | 10(4.4) | 7(3.1) | 6(2.6) |  | 0(0) | 1(2.8) | 0(0) | 3(8.3) |  | 4(2.9) | 7(5.1) | 5(3.7) | 1(0.7) |  |
| Severe | 0(0) | 4(1.8) | 2(0.9) | 6(2.6) |  | 0(0) | 0(0) | 0(0) | 2(5.6) |  | 0(0) | 0(0) | 3(2.2) | 3(2.2) |  |
|  | Post MR Severity |  |  |  |  | Post MR Severity |  |  |  |  | Post MR Severity |  |  |  |  |
| Pre-MR Severity | None | Mild | Moderate | Severe | p-value | None | Mild | Moderate | Severe | p-value | None | Mild | Moderate | Severe | p-value |
| None | 6(2.7) | 29(12.9) | 4(1.8) | 0(0) | <0.01* | 0(0) | 5(13.9) | 0(0) | 0(0) | 0.08 | 6(4.5) | 14(10.4) | 1(0.7) | 0(0) | <0.01* |
| Mild | 24(10.7) | 103(45.8) | 16(7.1) | 3(1.3) |  | 3(8.3) | 16(44.4) | 4(11.1) | 1(2.8) |  | 21(15.7) | 60(44.8) | 6(4.5) | 0(0) |  |
| Moderate | 1(0.4) | 15(6.7) | 17(7.6) | 1(0.4) |  | 0(0) | 1(2.8) | 4(11.1) | 1(2.8) |  | 1(0.7) | 14(10.4) | 7(5.2) | 0(0) |  |
| Severe | 0(0) | 2(0.9) | 2(0.9) | 2(0.9) |  | 0(0) | 0(0) | 1(2.8) | 0(0) |  | 0(0) | 2(1.5) | 2(1.5) | 0(0) |  |
|  | Pre |  | Post |  | p-value | Pre |  | Post |  | p-value | Pre |  | Post |  | p-value |
| LVEF, % | 62.5 [58.0, 66.0] |  | 59.0 [54.0, 63.3] |  | <0.01* | 60.1 ± 10.5 |  | 52.5 ± 14.4 |  | <0.01* | 59.5 [51.0, 64.0] |  | 58.5 [53.0, 65.0] |  | 0.35 |
| LVIDd, cm | 4.66 ± 0.66 |  | 4.63 ± 0.69 |  | 0.37 | 4.64 ± 0.84 |  | 4.75 ± 0.93 |  | 0.34 | 4.71 [4.26, 5.20] |  | 4.58 [4.29, 4.96] |  | 0.02* |
| RVIDd, cm | 3.85 ± 0.66 |  | 3.76 ± 0.66 |  | 0.18 | 3.98 ± 0.90 |  | 3.82 ± 0.86 |  | 0.35 | 3.79 ± 0.66 |  | 3.69 ± 0.61 |  | 0.02* |
| TAPSE, cm | 2.15 ± 0.51 |  | 2.03 ± 0.49 |  | <0.01* | 1.68 ± 0.51 |  | 1.74 ± 0.53 |  | 0.92 | 2.14 ± 0.49 |  | 2.06 ± 0.47 |  | 0.08 |

Data presented as number, n (%), median [25<sup>th</sup> percentile, 75<sup>th</sup> percentile] or mean ± SD. Comparisons were made using the chi-squared test for frequencies, and paired t-test or Wilcoxon signed-rank test for continuous variables.
Abbreviations: LBBA = left bundle branch area, LVEF = left ventricular ejection fraction, LVIDd = left ventricular internal diameter in diastole, MR = mitral regurgitation, RV = right ventricular, RVIDd = right ventricular internal diameter in diastole, TAPSE = tricuspid annular plane systolic excursion, TR = tricuspid regurgitation

#### Within patient changes

The median change in TR grade was 0 [0 to 1] in patients with conventional RV pacemakers, 0 [0 to 1] in patients with leadless pacemakers, and 0 [0 to 0] in patients with LBBA pacemakers and differed across pacemaker types (p<0.01 overall, LBBA vs leadless p<0.01, LBBA vs RV p=0.02, leadless vs RV p=0.51; Table 4). Figure 2 illustrates within patient change in TR grade for each pacemaker type.

**Figure 2.**
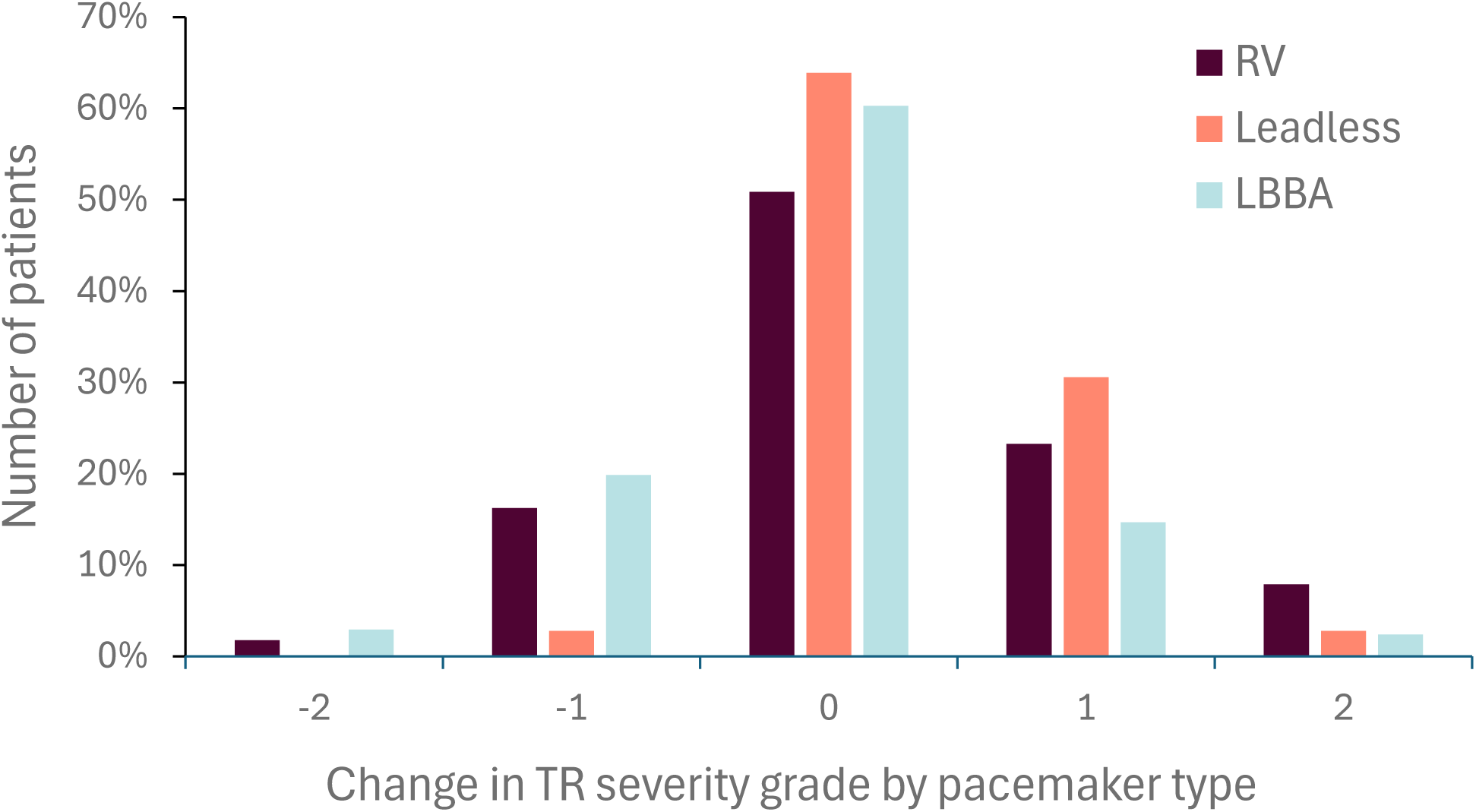
Within patient change in TR grade following each pacemaker type. The median change in TR grade differed across pacemaker types (p<0.01). Change in grade: improved by 2 grades (-2), improved by 1 grade (-1), no change in grade (0), worsened by 1 grade (1), worsened by 2 grades (2). Abbreviations: LBBA = left bundle branch area, RV = right ventricular, TR = tricuspid regurgitation

#### Prevalence of TR changes and regression models

The prevalence of severe TR was similar between groups at baseline, but lower after LBBA pacemaker implantation compared to both conventional RV and leadless pacemaker implantation (Table 3). The incidence of TR worsening by ≥ 1 grade was 26.5% overall and was more common following conventional RV compared to LBBA pacemakers (p<0.01 overall, LBBA vs RV p<0.01). The incidence of TR worsening by ≥ 2 grades did not differ between pacemaker groups (p=0.05 overall). In multivariable ordinal logistic regression, independent predictors of change in TR severity included conventional RV or leadless pacemaker type compared to LBBA and baseline TR severity (Supplemental Table 1). Leadless (OR 2.26, p=0.03) and RV pacemakers (OR 1.66, p=0.03) compared to LBBA predicted worsening TR, while lower TR at baseline (OR 0.33, p<0.01) predicted less decline in tricuspid valve function.

**Table 3.** Prevalence of severe TR and incidence of TR worsening after each pacemaker type.

|  | **RV**<br>**n=228** | **Leadless**<br>**n=36** | **LBBA**<br>**n=136** | **p-value,**<br>**overall** | **RV vs**<br>**Leadless,**<br>**p-value** | **RV vs**<br>**LBBA,**<br>**p-value** | **Leadless vs**<br>**LBBA, p-**<br>**value** |
| --- | --- | --- | --- | --- | --- | --- | --- |
| Severe TR, n (%) | 28 (12.3%) | 6 (16.7%) | 6 (4.4%) | **0.02\*** | 0.47 | **0.01\*** | **0.01\*** |
| TR worsening ≥1<br>grade, n (%) | 71 (31.1%) | 12 (33.3%) | 23 (16.9%) | **<0.01\*** | 0.79 | **<0.01\*** | 0.03 |
| TR worsening ≥2<br>grades, n (%) | 18 (7.9%) | 1 (2.8%) | 3 (2.2%) | 0.05 | 0.27 | 0.02 | 0.84 |

Data presented as number, n (%). Comparisons were made using the chi-squared test, with Bonferroni correction for multiple comparisons. \* $p < 0.05$ overall, \* $p < 0.05/3$ in pairwise comparisons.
Abbreviations: LBBA = left bundle branch area, RV = right ventricular, TR = tricuspid regurgitation

**Table 4.**
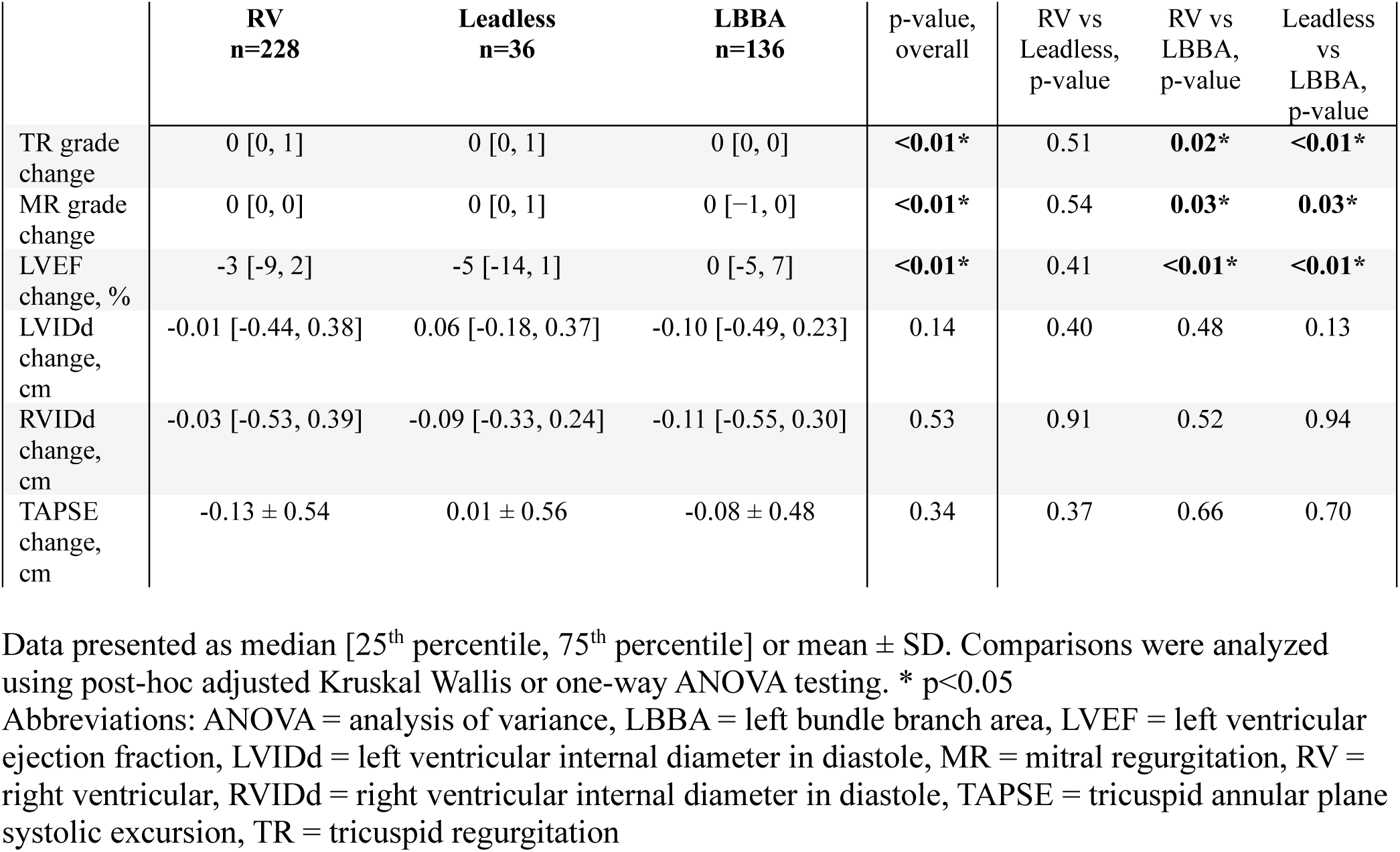
Within patient change in echocardiographic measurements compared between pacemaker types.

|  | **RV**<br>**n=228** | **Leadless**<br>**n=36** | **LBBA**<br>**n=136** | p-value,<br>overall | RV vs<br>Leadless,<br>p-value | RV vs<br>LBBA,<br>p-value | Leadless<br>vs<br>LBBA,<br>p-value |
| --- | --- | --- | --- | --- | --- | --- | --- |
| TR grade<br>change | 0 [0, 1] | 0 [0, 1] | 0 [0, 0] | **<0.01\*** | 0.51 | **0.02\*** | **<0.01\*** |
| MR grade<br>change | 0 [0, 0] | 0 [0, 1] | 0 [-1, 0] | **<0.01\*** | 0.54 | **0.03\*** | **0.03\*** |
| LVEF<br>change, % | -3 [-9, 2] | -5 [-14, 1] | 0 [-5, 7] | **<0.01\*** | 0.41 | **<0.01\*** | **<0.01\*** |
| LVIDd<br>change,<br>cm | -0.01 [-0.44, 0.38] | 0.06 [-0.18, 0.37] | -0.10 [-0.49, 0.23] | 0.14 | 0.40 | 0.48 | 0.13 |
| RVIDd<br>change,<br>cm | -0.03 [-0.53, 0.39] | -0.09 [-0.33, 0.24] | -0.11 [-0.55, 0.30] | 0.53 | 0.91 | 0.52 | 0.94 |
| TAPSE<br>change,<br>cm | -0.13 ± 0.54 | 0.01 ± 0.56 | -0.08 ± 0.48 | 0.34 | 0.37 | 0.66 | 0.70 |

Data presented as median [25<sup>th</sup> percentile, 75<sup>th</sup> percentile] or mean ± SD. Comparisons were analyzed using post-hoc adjusted Kruskal Wallis or one-way ANOVA testing. \* p<0.05
Abbreviations: ANOVA = analysis of variance, LBBA = left bundle branch area, LVEF = left ventricular ejection fraction, LVIDd = left ventricular internal diameter in diastole, MR = mitral regurgitation, RV = right ventricular, RVIDd = right ventricular internal diameter in diastole, TAPSE = tricuspid annular plane systolic excursion, TR = tricuspid regurgitation

### Changes in mitral valve function

#### Group level changes

The overall MR grade in the entire cohort was unchanged after pacemaker implantation (MR grade pre: 1 [1, 1], MR grade post: 1 [1, 1]; p=0.96). The frequency distribution of MR grade severity was different pre-implant compared to post-implant in RV and LBBA group, but not the leadless group (RV, p<0.01; leadless, p=0.08; LBBA, p<0.01).

#### Within patient changes

The median change in MR grade was 0 [0 to 0] in patients with conventional RV pacemakers, 0 [0 to 1] in patients with leadless pacemakers, and 0 [-1 to 0] in patients with LBBA pacemakers and differed across pacemaker types (p<0.01 overall, LBBA vs leadless p=0.03, LBBA vs RV p=0.03, leadless vs RV p=0.54; Table 4).

#### Regression models

In multivariable ordinal logistic regression, independent predictors of change in MR severity included conventional RV or leadless pacemaker type compared to LBBA, and baseline MR severity (Supplemental Table 2). Leadless (OR 3.43, p<0.01) and RV pacemakers (OR 1.84, p=0.01) compared to LBBA predicted worsening MR, while lower MR at baseline (OR 0.08, p<0.01) predicted less decline in mitral valve function.

### Changes in ventricular function and size

#### Group level changes

LVEF in the entire cohort decreased following pacemaker implantation (LVEF pre: 61.0 [57.0, 65.0]%, LVEF post: 59.0 [54.0, 64.0]%; p<0.01). There was a reduction in median LVEF post versus pre-implant in the conventional RV (p<0.01) and leadless (p<0.01) pacemaker groups but no change in the LBBA group (p=0.35, Table 2). Median LVIDd and RVIDd were reduced in the LBBA group (p=0.02 and p=0.02 respectively) but were unchanged in the conventional RV and leadless groups. TAPSE decreased post versus pre-implant in the conventional RV group (p<0.01) but did not change in the leadless and LBBA groups.

#### Within patient changes

LVEF worsened more in the conventional RV group (-3 [-9 to 2]%) and leadless group -5 [-14 to 1]% compared to the LBBA group 0 [-5 to 7]% (p<0.01 overall, LBBA vs leadless p<0.01, LBBA vs RV p<0.01, leadless vs RV p=0.41; Table 4). The within patient change of TAPSE, LVIDd, and RVIDd did not differ across the pacemaker types.

## Discussion

This study investigated the relationship between pacemaker type and various key echocardiographic measures of valvular and ventricular function following implantation of conventional transvenous RV, leadless RV, and LBBA pacemakers. The main findings were: 1) Over a median follow up of 15 months, TR worsened in one quarter of patients following pacemaker implantation. 2) Group level and within patient analyses demonstrated that TR and MR severity, and LVEF, worsened more in patients receiving conventional RV and leadless pacemakers compared to LBBA pacemakers. 3) In multivariable regression models, implantation of both RV or leadless pacemakers independently predicted worsening TR and MR, when compared to implantation of LBBA pacemakers.

### Changes in tricuspid valve function

The overall incidence of TR worsening by ≥1 grade was 26.5% and worsening by ≥2 grades was 5.5% following pacemaker implantation. A meta-analysis has shown a similar incidence of CIED-associated TR worsening ≥1 grade at 24%, but a slightly higher incidence of ≥2 grades at 11% (13). An explanation for the lower incidence observed in this study could be that the meta-analysis included additional types of CIEDs including cardiac resynchronization therapy devices and implantable cardioverter defibrillators. Furthermore, conventional RV pacemakers which had a higher incidence of TR worsening in our study, likely formed a greater portion of the permanent pacemakers included in the meta-analysis. The current study found a similar rate of worsening by ≥2 grades of 7.9% among patients receiving conventional RV pacemakers. Our study may have overestimated the incidence of post-pacemaker TR, by selecting the post-implant echocardiogram with the most severe TR grade and longest follow-up. This echocardiogram was chosen to avoid missing cases of subsequent severe TR, which can be underestimated with echocardiographic assessment (14, 15).

This study observed worsening TR severity following conventional RV and leadless pacemaker implantation but not following LBBA pacemaker implantation. Previous studies investigating change in TR severity after pacemaker implantation have yielded inconsistent results. Most studies show TR worsening after RV pacing (4, 16). Findings after leadless pacemaker implant have been contradictory with some studies reporting worsening TR (17, 18) and others reporting no change (9, 19). Some studies have shown TR improvement after CSP (20, 21) while others have shown TR worsening (22, 23). Prior studies of TR after CSP primarily included patients with HBP and few patients with LBBA. This study was performed to better characterize changes in TR severity in the largest cohort of LBBA pacemakers reported to date. Further work in other multicenter patient cohorts is needed to further clarify if pacemaker type affects TR development.

The mechanisms contributing to TR worsening after pacemaker implantation are likely complex. Among patients receiving LBBA pacemakers, mechanical lead-valve interactions may be the dominant mechanism. Previous studies have found lead placement further from the tricuspid valve associated with lower TR grade after implant (22, 23). The observed increased risk of worsening TR among patients receiving conventional or leadless RV pacemakers may be due to pacing induced ventricular dyssynchrony and subsequent functional TR, which may be exacerbated by non-physiologic pacing and alleviated by physiologic pacing. Interestingly, leadless pacemakers were associated with a higher odds ratio for predicting worsening TR compared to RV pacemakers. Despite using a similar pacing mechanism, this difference could be due to leadless pacemakers having a bigger disturbance on subvalvular tricuspid structures during and following implantation. This is a novel finding and needs further investigation, but could alter pacemaker type selection for patients with an imminent TR risk. Additionally, greater baseline TR grade also independently predicted worsening TR, an observation which contributes to the known risk factors for CIED-related TR (24).

Despite similar prevalence of severe TR between pacemaker groups before implant, progression to severe TR was more frequent after RV and leadless implantation compared to LBBA implant. To our knowledge this is the first study comparing this clinically relevant endpoint following different pacemaker types. Previous studies have reported a varying prevalence of severe TR after pacemaker implant between 3%-12% (25–27) and worsening ≥2 grades between 1-18% (28). However, most of these studies included multiple CIED types without distinguishing between different pacemaker types. The differences in severe TR frequencies we observed following RV and LBBA pacemaker implant needs further validation, but may carry clinical implications. Severe TR is associated with increased heart failure symptom severity, heart failure hospitalizations and reduced time to death (5, 29).

### Changes in mitral valve function

In the current study, MR grade improved more following LBBA, compared to RV and leadless pacemaker implantation. Prior studies have shown different outcomes on MR and did not include comparisons between pacemaker types (6, 17, 19, 30). However, Yuyun et al. (4) conducted a meta-analysis comparing the prevalence of ≥ moderate MR before and after pacemaker implantation. They concluded that the risk of deterioration in MR increased after RV, decreased after CSP, and remained unaltered after leadless pacemaker. Interestingly, the meta-analysis further described that CRT also reduced the risk of post-device MR in a manner similar to CSP. This may be due to the benefits on ventricular function that both CSP and CRT have compared to RV pacing (7), and could signify that the MR improvement may be caused by the improvement in global ventricular synchrony and LV function. Additionally, in contrast to TR, higher RV pacing percentage is associated with worsening MR (31), which strengthens this reasoning. Similarly to TR, independent predictors of worsening MR were implantation of leadless and RV pacemakers compared to LBBA pacemakers, and a greater baseline MR grade. Correspondingly to the findings previously discussed, these findings could have an impact on device selection strategies.

### Changes in ventricular function and size

Worsening LV function after RV pacemaker implantation, or pacing induced cardiomyopathy (PICM), occurs in 12-25% of patients, and may be mitigated by CSP (32–34). Because leadless pacemakers currently also pace the RV myocardium, they may also cause PICM (19, 35). Our study showed a reduction in LVEF following implant of RV and leadless pacemakers compared to LBBA pacemakers, with no difference in LVEF change between conventional RV and leadless RV pacemakers, which has also been described in previous studies (19).

RV pacemakers have been shown to be associated with a reduction in right ventricular ejection fraction (RVEF), with a similar reduction in other RV function measurements, such as fractional area change (FAC), TAPSE and S’ (peak systolic) velocity, 6 months after RV pacemaker implantation (36). Further in the same study, multiple RV and right atrial dimensions were measured with only right ventricular outflow tract proximal diameter increasing, and the others remaining unchanged. In the current study there was a decrease in TAPSE post versus pre-implant following RV pacemakers but not after the other pacemaker types. A decrease in right ventricular systolic function could contribute to the observed worsening TR in the RV pacemaker group. However, no difference in within patient change in TAPSE was found between pacemaker types and baseline TAPSE did not predict change in TR, MR or LVEF.

### Limitations

This was a single-center observational study, and as such the external validity may be limited. Second, the retrospective study design prompts various limitations related to selection bias due to echocardiograms before and after pacemaker implantation being performed at physicians’ discretion, non-randomization of pacemaker groups, and missing echocardiographic data. Third, the leadless group was substantially smaller compared to the other pacemaker groups, due to infrequent leadless implantations in the study timeline. Finally, this study did not utilize three-dimensional echocardiography to delineate lead and valve interactions, and a causal relationship between pacemaker implant and the echocardiographic changes described in this study cannot be confirmed.

## Conclusion

Overall, TR worsened, MR remained unchanged and LVEF worsened following pacemaker implantation. LBBA pacemakers displayed more favorable outcomes on tricuspid and mitral valve function, and left ventricular function compared to RV and leadless pacemakers. Furthermore, severe TR was more common following implantation of RV and leadless pacemakers compared to LBBA pacemakers.

## Data availability statement

The data that supports the findings of this study are available on request from the corresponding author. The data are not publicly available due to privacy or ethical restrictions.

## Ethics approval statement

An approval was obtained by the Inova Health System Institutional Review Board (IRB) with a retrospective waiver of individual informed consent (approval number INOVA-2024-86).

---

**Supplementary Table 1.** Predictive variables of ΔTR grade.

| Variables | Univariable analysis |  | Multivariable analysis |  |
| --- | --- | --- | --- | --- |
|  | OR (95% CI) | p-value | OR (95% CI) | p-value |
| CRT indication | 0.36 (0.14 to 0.97) | **0.04\*** | 0.36 (0.12 to 1.03) | 0.06 |
| ACE inhibitors /ARB/ARNI | 1.81 (1.24 to 2.66) | **<0.01\*** | 1.49 (0.99 to 2.23) | 0.05 |
| SGLT2 inhibitors | 2.59 (0.89 to 7.43) | 0.08 | 2.46 (0.81 to 7.42) | 0.11 |
| Months follow-up | 1.02 (1.00 to 1.04) | **0.03\*** | 1.02 (1.00 to 1.04) | 0.07 |
| Pacemaker type |  |  |  |  |
| Leadless vs LBBA | 2.59 (1.32 to 5.07) | **<0.01\*** | 2.26 (1.10 to 4.66) | **0.03\*** |
| RV vs LBBA | 1.81 (1.20 to 2.75) | **<0.01\*** | 1.66 (1.04 to 2.64) | **0.03\*** |
| Baseline TR grade | 0.31 (0.24 to 0.41) | **<0.01\*** | 0.33 (0.25 to 0.44) | **<0.01\*** |

Data presented as odds ratio (OR) and 95% confidence interval (95% CI). Variables $p < 0.1$ were included in the multivariable ordinal logistic regression model. \* $p < 0.05$ . Additional variables analysed in univariable regression that had $p > 0.1$ included age, sex, body surface area, hypertension, coronary artery disease, heart failure, atrial arrhythmias, chronic obstructive pulmonary disease, diabetes type 2, renal dysfunction, sinus node dysfunction, atrioventricular block, beta-blockers, mineralocorticoid receptor antagonists, loop diuretics, ventricular pacing percentage, baseline left ventricular ejection fraction, left ventricular internal diastolic dimension, right ventricular interval diastolic dimension, TAPSE and baseline MR grade.
Abbreviations: ACE = angiotensin-converting enzyme, ARB = angiotensin receptor blockers, ARNI = angiotensin receptor-neprilysin inhibitors, CRT = cardiac resynchronization therapy, LBBA = left bundle branch area, OR = odds ratio, SGLT2 = sodium-glucose cotransporter-2, TR = tricuspid regurgitation

**Supplementary Table 2.** Predictive variables of ΔMR grade.

| Variables | Univariable analysis |  | Multivariable analysis |  |
| --- | --- | --- | --- | --- |
|  | OR (95% CI) | p-value | OR (95% CI) | p-value |
| CRT indication | 2.37 (0.86 to 6.60) | 0.10 | 0.61 (0.18 to 2.16) | 0.43 |
| Body surface area | 0.50 (0.24 to 1.06) | 0.07 | 0.84 (0.36 to 1.99) | 0.70 |
| Pacemaker type |  |  |  |  |
| Leadless vs LBBA | 0.40 (0.20 to 0.82) | **0.01\*** | 3.43 (1.57 to 7.52) | **<0.01\*** |
| RV vs LBBA | 0.58 (0.38 to 0.88) | **0.01\*** | 1.84 (1.15 to 2.97) | **0.01\*** |
| Baseline TR grade | 1.47 (1.13 to 1.90) | **<0.01\*** | 1.22 (0.90 to 1.65) | 0.19 |
| Baseline MR grade | 10.5 (7.25 to 15.4) | **<0.01\*** | 0.08 (0.05 to 0.13) | **<0.01\*** |
| Baseline LVEF | 0.98 (0.96 to 1.00) | 0.05 | 1.00 (0.98 to 1.02) | 0.87 |

Data presented as odds ratio (OR) and 95% confidence interval (95% CI). Variables $p < 0.1$ were included in the multivariable ordinal logistic regression model. \* $p < 0.05$ . Additional variables analysed in univariable regression that had $p > 0.1$ included age, sex, hypertension, coronary artery disease, heart failure, atrial arrhythmias, chronic obstructive pulmonary disease, diabetes type 2, renal dysfunction, sinus node dysfunction, atrioventricular block, angiotensin-converting enzyme/angiotensin receptor blockers/angiotensin receptor-neprilysin inhibitors, beta-blockers, mineralocorticoid receptor antagonists, loop diuretics, sodium-glucose cotransporter-2 inhibitors, ventricular pacing percentage, baseline left ventricular internal diastolic dimension, right ventricular interval diastolic dimension and TAPSE. Abbreviations: ACE = angiotensin-converting enzyme, ARB = angiotensin receptor blockers, ARNI = angiotensin receptor-neprilysin inhibitors, CRT = cardiac resynchronization therapy, LBBA = left bundle branch area, OR = odds ratio, TR = tricuspid regurgitation, MR = mitral regurgitation

